# Status of physical activity and its associated factors among the secondary school teachers in Pokhara, Nepal

**DOI:** 10.1101/2024.01.04.24300849

**Authors:** Kamal Ranabhat, Shubhadra Shahi, Kiran Shrestha, Ramesh Kunwar, Himalaya Rana, Bishnu Prasad Choulagai

**Affiliations:** Central Department of Public Health, Institute of Medicine, Tribhuwan University, Kathmandu, Nepal; Ministry of Health and Population, Bagmati Province, Nepal

## Abstract

Cardiovascular diseases are the leading cause of all the cause of death globally and physical activity is the proven cost effective modifiable behavior risk factors for the prevention of cardiovascular diseases. The objective of the study was to assess the status of the physical activity and its determining factors among the school teachers of Pokhara Nepal. We used quantitative approach for data collection among 406 samples that were drawn using probability proportion to enrollment size. The international questionnaire on physical activity (IPAQ)-long form was used for data collection. SPSS V.27 was used for univariate and bivariate analysis.

The study have found the 13.2 percent of prevalence of physical inactivity. Sex, ethnicity, educational qualification, availability of walking environment around home, mean screen time per day were found having statistical association with the outcome variable. The prevalence of low physical activity was found 13.2 percent. Majority of the study participants achieved WHO global recommendation on physical activity. Domestic and garden work contributed most in domain specific physical activity among the participants.

## Introduction

Cardiovascular diseases are a set of disorders related to heart and blood vessels, which includes coronary heart disease, cerebrovascular disease, peripheral arterial disease, rheumatic heart disease, congenital heart disease and thrombosis of deep vein and pulmonary embolism. Heart attacks and strokes, which cause by blockage preventing flow of blood to heart or brain, are frequently used to generalize all heart disorders ^[1]^.

It is a leading cause of death globally killing estimated 17.9 million life in 2019, which represents almost 32% of all global deaths. The burden of cardiovascular disease can be reflected from the evidence that of 17 million premature mortality caused by non-communicable diseases, 38% are attributed to cardiovascular diseases ^[2,3]^.

The world scenario suggest that there is a constant increase in the CVDs related deaths around the world. The disease which accounted 17.5 million deaths in 2012 rocketed to 17.9 million death in 2016 and with this rate of inclination, by 2030, it is estimated that 22.2 million deaths will be attributed to CVDs ^[3]^. Nearly half of the burden can be prevented with lifestyle modification such as physical activity, eating behavior, altering drinking and smoking habits ^[2,3]^.

The status of performing physical activity globally is below WHO recommendations and in Nepal also the prevalence of physical inactivity has inclined from 2.4% to 7.4% between 2013 and 2019 among adults of 18-69 years^[4,5]^. The study conducted among 40-80 years respondents in Lumjung district in 2014 shows the prevalence of low physical activity of 10% ^[6]^.

Moreover, the unnecessary direct health care cost to physical inactivity is $54 billion annually and additional $14 billion cost on compromised productivity. Thus, the global action plan on physical activity have set an ambitious target of 15% reduction in physical inactivity prevalence by 2030 ^[5]^.

Regular physical activity is the proven cost effective intervention of cardiovascular diseases ^[7,8]^. There are several ways physical activities can be performed such as cycling, walking, engaging in sports and other forms of recreations like yoga and dance ^[5]^. As per the WHO recommendation, moderate physical activity of at least 150 minutes or vigorous intensity physical activity of 75 minutes or combination of both is required for healthy life ^[5,7]^. The study aims to assess the status of physical activity and its associated factors among school teachers of Pokhara, Nepal.

## Methods and materials

### Study design and population

This study was school-based descriptive cross-sectional study conducted in secondary schools of Pokhara metropolitan city. The probability proportionate to enrollment size sampling technique was used for obtaining the required sample size. The list of all the secondary level schools and no. of school teachers was obtained from Pokhara metropolitan city. There are 111 secondary schools and 2044 school teachers in Pokhara metropolitan city. With reference to the 80% prevalence of the physical inactivity among the civil servant in Nepal ^[9]^, at 5% allowable error, 5% level of significance and 1.5 times design effect, the required sample size was 406. The school response rate was 90.91% and participant’s response rate was 87.58%.

### Study variables and data collection tools

#### Outcome variables

The outcome variable of the study is status of physical activity. Participant status of the physical activity was assessed using IPAQ-long form. The physical activity status was measured in four domains specific (work, transport, domestic/garden and leisure time) and three activity specific physical activity (walking, moderate and vigorous physical activity) in a week. The status of the physical activity was classified as high level, moderate level and low level of physical activity.

##### High level physical activity

Participants who meets any of the following criteria was considered as having high level physical activity;

Vigorous-intensity activity on at least 3 days achieving a minimum total physical activity of at least 1500 MET-minutes/week or

7 or more days of any combination of walking, moderate-intensity or vigorous-intensity activities achieving a minimum total physical activity of at least 3000 MET-minutes/week.

##### Moderate level physical activity

Meeting any of the condition below was considered as moderate level physical activity

3 or more days of vigorous-intensity activity of at least 20 minutes per day or

5 or more days of moderate-intensity activity and/or walking of at least 30 minutes per day **or**

5 or more days of any combination of walking, moderate-intensity or vigorous intensity activities achieving a minimum total physical activity of at least 600 MET-minutes/week.

##### Low level physical activity

participants who fail to meet the criteria for moderate or high level was considered to have low level physical activity.

#### Independent variable

Through the relevant literature review, independent variables were carefully selected and classified in the different thematic categories such as demographic variables, socioeconomic variables, environmental variables and existing knowledge and practice on physical activity.

Demographic variables included age, sex, marital status, family type, ethnicity. Socio-economic variable included education. Environmental variable included availability of open space (park), gym hall, walking space around house.

### Statistical analysis

Data were entered and analyzed using Statistical Package for Social Science (SPSS v27.0). Data entry was cautiously done to minimize the errors.

Descriptive statistics such as frequency, percentage, mean and standard deviation was calculated using univariate analysis while bivariate analysis was done to test the existence of the significant association between dependent and independent variables and for assessing the weighted odds ratio between dependent and independent variables and determining the factors associated with physical activity, multiple logistic regression analysis was performed.

The multi-collinearity test shown none of the variable having tolerance less than 0.1 and variance inflation factor was observed less than 10 suggesting no association or interdependence between the different independent variables. The p-value was less than 0.05 implying that the model is statistically significant and predicts the dependent variables.

### Ethical consideration

Ethical approval was obtained from the Institutional Review Committee (IRC) of the Institute of Medicine, Maharajgunj [Ref:259(6-11)E2 2079/080]. Similarly, the formal permission was also obtained from the Pokhara metropolitan city and respective schools. Prior to data collection, the objective and methodology of the study was clearly stated to the participants. The voluntary participation was encouraged and also the participant withdrawal from the study at any time was respected. Privacy, confidentiality and anonymity was ensured. The written informed consent signed by participants who were willing to take part in the study was considered for data collection.

## Results

### Socio-demographic characteristics of the study participants

Of the study respondents, 64.8 percent were from private school while remaining 35.2 percent were from the public school and 53 percent were male. Ethnicity wise, Brahmin and Chhetri were among the major ethnic group (69.2%), followed by Janajati (21.3%). Very few study participants were among Dalit (3.4%), Sanyasi/Thakuri (3.4%) and Madeshi (1.2%).

Similarly, 21.5 percent were among 20-30 years of age group, 42.3 percent were found among the age group of 31-40 years, 24.0 percent were found among the age group of 41-50 years while only 12.2 percent of the study participants were above 50 years of age. The mean age of the study participants was found 38.65 years with standard deviation 9.469 while mean years of teaching was found 15.58 years with standard deviation 12.025.

Similarly, more than half (53.3%) study participants had master degree followed by bachelor degree (31.5%) and higher secondary (11.2%). Majority of the study participants (82.2%) had married marital status while 17.8 percent were found unmarried. More than half of the study participants used to live in joint and extended family followed nuclear family type.

**Table 1.** Distribution of the participants by socio-demographic characteristics (n=409) Characteristics.

| **Characteristics** |  | **Number** | **Percent** |
| --- | --- | --- | --- |
| **School type** | Public | 144 | 35.2 |
|  | Private | 265 | 64.8 |
| **Sex** | Male | 217 | 53 |
|  | Female | 192 | 47 |
| **Ethnicity** | Dalit | 14 | 3.4 |
|  | Janajati | 87 | 21.3 |
|  | Bhramin/Chhetri | 283 | 69.2 |
|  | Others | 25 | 6.2 |
| **Respondents age** | 20-30 Years | 88 | 21.5 |
|  | 31-40 Years | 173 | 42.3 |
|  | 41-50 Years | 98 | 24.0 |
|  | ≥51 Years | 50 | 12.2 |
| **Mean age of the participants (SD)** |  | **38.65(9.469)** |  |
| **Years of Teaching** | Upto 5 Years | 67 | 16.4 |
|  | 6-10 Years | 81 | 19.8 |
|  | 11-15 Years | 95 | 23.2 |
|  | 16-20 Years | 70 | 17.1 |
|  | 21 and above | 96 | 23.5 |
| **Mean Years of teaching (SD)** |  | **15.58 (12.025)** |  |
| **Educational qualification** | Higher secondary | 46 | 11.2 |
|  | Bachelor | 129 | 31.5 |
|  | Masters | 218 | 53.3 |
|  | MPhil | 9 | 2.2 |
|  | PhD | 7 | 1.7 |
| **Marital Status** | Married | 336 | 82.2 |
|  | Unmarried | 73 | 17.8 |
| **Family type** | Nuclear | 189 | 46.2 |
|  | Joint/Extended | 220 | 53.8 |

### Physical activity status

Among the study participants, low level of physical activity was found in 13.2 percent. The moderate level of physical activity and high level of physical activity was found in 65.3 and 21.5 percent respectively.

Study showed that 86.3 percent of the total respondents meeting the WHO global recommendation on physical activity.

**Table 2:** Status of physical activity.

**n=409**
| Characteristics | Number | Percent | 95% CI |
| --- | --- | --- | --- |
| **IPAQ classification** |  |  |  |
| Low Physical Activity | 54 | 13.2 | 0.10-0.17 |
| Moderate Physical Activity | 267 | 65.3 | 0.61-0.70 |
| High Physical Activity | 88 | 21.5 | 0.18-0.26 |
| WHO global recommendation on physical activity | 355 | 86.3 | 0.84-0.90 |

### Activity and domain specific physical activity

Activity specific analysis shows moderate activity (52.36%) as the highest contributor of total MET-minutes/week followed by walking (26.25%) and vigorous activity (21.38%). Domain wise analysis shows that, maximum contributor of MET-minute/week is domestic and garden work (34.39%), followed by work (31.38%), leisure time activity (21.89%) and transport (12.34%).

**Table 3:** Activity and domain specific physical activity.

**n=409**
| Total PA score | Total (%) | Median MET-minutes/week | IQR |
| --- | --- | --- | --- |
|  |  | 3561 | 1324~6733 |
| **Activity specific** |  |  |  |
| Walking | 26.25 | 780 | 219~1647 |
| Moderate | 52.36 | 1665 | 462.50~3780 |
| Vigorous | 21.38 | 240 | NA |
| **Domain Specific** |  |  |  |
| Work | 31.38 | 840 | 230~2020 |
| Transport | 12.34 | 247 | NA |
| Domestic and Garden | 34.38 | 870 | 180~2520 |
| Leisure Time | 21.89 | 438 | 63~1320 |

### Enabling environment for the physical activity

Among the study participants, 59.2 percent reported the availability of play ground around their home environment. Almost 82 percent study participants reported having open space for walking near their home. Similarly, most of them (64.5%) reported having physical activity facilities around home. Availability of football or volleyball ground and badminton courts was the common type of physical activity facilities around their residential areas. In order to engage in physical activities beside job, most of the participants (67% and 69.2%) had farming land and kitchen garden respectively. Study participants also reported of animal husbandry (24.7%), cycle (37.6%), garden (45.7%) where participants could engage in different level of physical activities.

Moreover, motorcycle (61.4%) was the most common means of transport used by the study participants followed by public bus (13.2%). Very few participants used cycle and private car as a means of transportation for daily travel to job. Similarly, majority of the teachers (87.8%) reported that they usually teach by walking around the classroom while only few teachers (10.8%) reported that they usually sit in a place while teaching and very negligible (1.5%) teachers reported that their teaching way was standing position.

Study found that, only one fourth of the study participants had to travel more than 30 minutes to reach school while more than three fourth (76%) had to walk less than 30 minutes to reach school from home.

**Table 4:** Enabling environment for the physical activity.

**n=409**
| **Characteristics** |  | **Number** | **Percent** |
| --- | --- | --- | --- |
| **Availability of play ground around home** | Yes | 242 | 59.2 |
|  | No | 167 | 40.8 |
| **Availability of open space for walking** | Yes | 334 | 81.7 |
|  | No | 75 | 18.3 |
| **Physical activity facility around home** | Yes | 264 | 64.5 |
|  | No | 145 | 35.5 |
| **Physical activity facility around home** | Gym | 146 | 55.3 |
|  | Football/Volleyball ground | 172 | 65.2 |
|  | Cricket | 43 | 16.3 |
|  | Swimming Pool | 73 | 27.7 |
|  | Badminton | 156 | 59.1 |
|  | Table Tanis | 61 | 23.1 |
| **Physical activity facility outside job** | Yes | 279 | 68.2 |
|  | No | 130 | 31.8 |
| **Engaging in physical activity beside job** | Farming land (Khetbari) | 187 | 67.0 |
|  | Animal husbandry | 69 | 24.7 |
|  | Physical activity stools | 87 | 31.2 |
|  | Cycle | 105 | 37.6 |
|  | Kitchen garden | 193 | 69.2 |
|  | Garden | 128 | 45.7 |
| **Usual Means of Transport to school** | Walking | 90 | 22.0 |
|  | Cycle | 10 | 2.4 |
|  | Motorcycle | 251 | 61.4 |
|  | Public Bus | 54 | 13.2 |
|  | Car | 4 | 1.0 |
| **Usual way of teaching** | Sitting | 44 | 10.8 |
|  | Standing | 6 | 1.5 |
|  | Walking | 359 | 87.8 |
| **Time taken to reach school from home** | ≤30 minutes | 311 | 76.0 |
|  | >30 minutes | 98 | 24.0 |

### Source of motivation for the physical activity

Of the total, respondents, almost half (45%) received the motivation to physical exercise from the media, while the adverse health conditions of family members or friends due to lack of physical exercise and experts were also the common source of motivation for the respondents to be engaged in physical exercise. Very few (6.1%) of the respondents were motivated to engage in physical exercise due to deteriorating self health condition.

Similarly, social media (35.82%) was found to be the major source of information for the physical exercise among the study participants followed by suggestion and information from friends and colleague (15.71%), Television (12.66%), Newspaper (10.84%), Family (9.38%) and health workers (8.59%) while 7% respondents reported radio as the source information for physical activities.

**Table 5.**
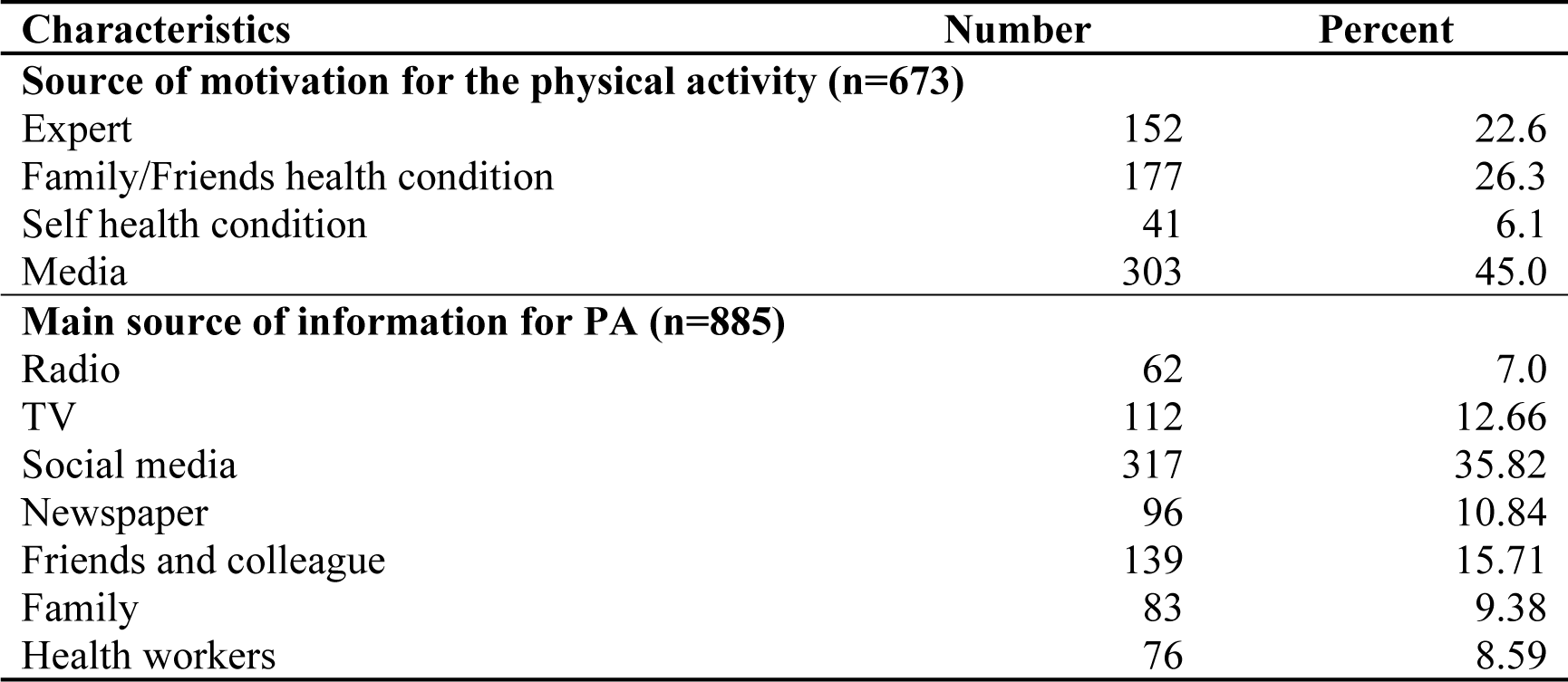
: Source of motivation for the physical activity.

| **Characteristics** | **Number** | **Percent** |
| --- | --- | --- |
| **Source of motivation for the physical activity (n=673)** |  |  |
| Expert | 152 | 22.6 |
| Family/Friends health condition | 177 | 26.3 |
| Self health condition | 41 | 6.1 |
| Media | 303 | 45.0 |
| **Main source of information for PA (n=885)** |  |  |
| Radio | 62 | 7.0 |
| TV | 112 | 12.66 |
| Social media | 317 | 35.82 |
| Newspaper | 96 | 10.84 |
| Friends and colleague | 139 | 15.71 |
| Family | 83 | 9.38 |
| Health workers | 76 | 8.59 |

### Sedentary Behavior

Sedentary behavior were assessed using the two variables i.e. average sitting time per day and average screen time per day.

The sitting time category for the study participants were categories as having sitting time upto 480 minutes per day and having sitting time above 480 minutes per day. Majority of the study participants had sitting time less than 480 minutes per day while remaining 10.5 percent had sitting time above 480 minutes per day. The mean sitting time for the respondents was found 273 minutes per day with SD 178.41.

Similarly, the mean screen time per day was found 205.33 minutes with 130.27 standard deviation. More teachers (56.7%) were found having screen time upto 180 minutes per day while remaining had more than 180 minutes per day screen time.

**Table 6:** Average sitting time and screen time per day.

**n=409**
| Characteristics | Number | Percent |
| --- | --- | --- |
| Average sitting time per day |  |  |
| ≤480 minutes | 366 | 89.5 |
| >480 minutes | 34 | 10.5 |
| Mean sitting time (SD) | 273 (178.41) |  |
| Average Screen time per day |  |  |
| ≤180 minutes | 232 | 56.7 |
| >180 minutes | 177 | 43.3 |
| Mean screen time (SD) | 205.33 (130.27) |  |

### Factors associated with physical activity

This table shows the logistic regression analysis of the variables having found statistical significant association in Chi-square test. Both unadjusted and adjusted logistic regression model for physical activity level is calculated using bivariate and multivariate logistic regression. The adjusted odds ratio shows that male were almost 2.193 folds of physically active compared to female counterpart. Similarly, participants with higher educational qualifications were also 2.88 times more engaged in moderate or high level of physical activity compared to participants with higher secondary and bachelor level of education. Compared to sitting and standing way of teaching, walking as a way of teaching was found almost 10 folds of engagement in moderate or high level of physical activity. Similarly, respondents having walkable area around their home were more likely to engage in regular physical activity compared to respondents having no walking area available around their home.

Ethnicity and mean screen time per day were found having significant association in bivariate logistic regression. However, in multiple logistic regression, they were not found significantly associated with physical activity.

**Table 11:** Factors associated with physical activity.

**n=409**
| Characteristics | Physical activity level |  |  |  |
| --- | --- | --- | --- | --- |
|  | Unadjusted OR<br>(95% CI) | P-value | Adjusted OR<br>(95% CI) | P-value |
| **Sex** |  |  |  |  |
| Male | **1.937 (1.08-3.48)** | **0.027\*** | **2.193(1.11-4.35)** | **0.025\*** |
| Female | **(ref)** |  | **(ref)** |  |
| **Ethnicity** |  |  |  |  |
| Janajati and other | 0.551 (0.31-0.99) | **0.046\*** | 1.064(0.53-2.13) | 0.861 |
| Bhramin/Chhetri | **(ref)** |  | **(ref)** |  |
| **Education qualification** |  |  |  |  |
| Master/MPhil/PhD | 2.892 (1.56-5.14) | **0.001\*** | 2.88(1.45-5.73) | **0.003\*** |
| Higher sec and bachelor | **(ref)** |  | **(ref)** |  |
| **Usual teaching way** |  |  |  |  |
| Walking | 10.12(5.19-19.76) | **<0.001\*** | 9.13(4.20-19.87) | **<0.001\*** |
| Sitting and standing | **(ref)** |  | **(ref)** |  |
| **Screen time per day** |  |  |  |  |
| >180 minutes | 0.525 (0.30-0.94) | **0.029\*** | 0.78(0.40-1.53) | 0.469 |
| ≤181 minutes | **(ref)** |  | **(ref)** |  |
| **Walking environment available around home** |  |  |  |  |
| Available | 3.44(1.88-6.29) | **<0.001\*** | 2.28(1.12-4.62) | **0.023\*** |
| Not available | **(ref)** |  | **(ref)** |  |

## Discussion

This study identified the prevalence of low physical activity 13.2 percent, moderate physical activity 65.3 percent and high level physical activity 21.5 percent. Reported prevalence of low physical activity in this study (13.2%) is high when compared to nationwide prevalence of low physical activity as per the STEP survey report finding (7.4%) ^[4]^. The prevalence of high physical inactivity in this study is supported by the argument that physical inactivity is more accumulated in the urban areas due to urban lifestyle as this study was also conducted in the urban setting ^[10]^. However, the global prevalence of the physical inactivity (27.7%) is almost double compared to the finding from this study ^[11]^, which may be due to the variation of the age group of the study population and also due to difference in the tool and technique used in the study.

While the prevalence of low physical inactivity of the study conducted in different place and time have shown almost 2 to 3 folds of high prevalence of low physical inactivity compared to current study ^[12–15]^. The time difference could be one reason as back in 2009, there was less awareness level about the importance of physical activity compared to today’s world where individual can acquire information from different sources. Also very fluctuating prevalence of low physical inactivity from different studies could be because of the use of different physical activity measuring tools.

The prevalence of low physical activity in current study is higher among female teachers compared to male counterparts which is similar to the other studies conducted in Iran and Brazilian teachers ^[16,17]^. However, the sex-wise prevalence contradicts with the study conducted in Chandigarh, which shows male teachers had the higher prevalence of low physical inactivity compared to female counterparts^[15]^.

In this study, domestic and garden related work domain contributed to the most in total MET-minutes per week which is conflicting with the finding of the NCD STEPS survey 2019 which shows that work domain contributed most in the total physical activity^[4]^. As Nepal is agrarian country and many teachers might be regularly involved in domestic chores such as working in kitchen garden, agricultural fields, animal husbandry and so on in the morning and evening, despite the fact that they are having a teaching job during day time.

The mean age of the teachers in this study is 38.65 with standard deviation ±9.47 which is almost similar to the study conducted in Brazil and Libya which shows the mean age of teachers 40 years and 36.96±8.63 years respectively ^[13,18]^.

Similarly, the existence of significant association of physical activity with qualification and availability of walking areas around house is consistence with the other studies ^[19,20]^. In the same way, the years of teaching have no significant association with the physical activity, which is supported by another study as well ^[19]^.

Another study conducted using GPAQ showed 97 percent of men and 98 percent of women met the WHO global recommendation level of physical activity, which is fairly high compared to current study, which shows only 86.6 percent study participants meeting the WHO global recommendation level for physical activity. However, another study shows equivalent status of meeting WHO global recommendation on physical activity ^[12,21]^.

The median MET-minute/week shown by current study is 3561 MET-minute/week, which is almost half the compared to another study which shows 8400 and 7140 MET-minutes/week separately for male and female respectively. Moreover, same study also shown that, work domain contributed to major energy expenditure contradicting with the finding from current study, which shows that domestic and garden domain contributed to most energy expenditure ^[21]^.

This study have shown statistical significant association between physical activity and other socio-demographic characteristics like sex, education qualifications, availability of walkable area near home and the way of teaching in classroom. Concurring with this finding, another report from Australia also have shown that availability of physical activity facilities having significant association with performing regular physical activity ^[22]^.

Reliable studies have shown sitting more than 8 hours per day is almost 32% likely to die of cardiovascular diseases compared to sitting less than 8 hours per day ^[23,24]^. Compared with this category of sitting time, 10.5 percent of the study participants were found having higher risk of cardiovascular diseases as they reported more than 8 hours per day of sitting time. Similarly, the mean sitting time of current study is found to be 273 minutes with 178.41 SD. This is higher compared to the mean occupational sitting time reported by another study conducted among adults in Australia ^[25]^. This could be because of the difference in the occupation type of the study participants. School teachers by their job nature, spend time in classroom either standing or walking. However, in the office and leisure time, they are engaged in table work in office contributing to more sitting time.

## Conclusions

The prevalence of low physical activity was found 13.2 percent and majority of the participants lie under the category of moderate physical activity. Almost 86.80% of the study participants achieved WHO global recommendation on physical activity. Domestic and garden work contributed most in domain specific physical activity among the participants. Female were more physically inactive compared to male counterparts. Sex, ethnicity, educational qualification, mean screen time, availability of walking area around home are some of the socio-demographic and other characters which were found having statistical significant association with the level of physical activity.

## Data Availability

All relevant data are within the manuscript and its Supporting Information files.

## Acknowledgement

It gives me immense pleasure to acknowledge all those who have given consistent guidance, advice and encouragement in my endeavor. I fully owe a favor to my academe Central Department of Public Health, IoM TU.

My regards and gratitude to all the Pokhara metropolitan city for timely granting the permission and school administration for managing time for data collection despite their busy schedule.

## Authors Contributions

Conceptualization: Kamal Ranabhat, Shubhadra Shahi, Kiran Shrestha, Himalaya Rana, Rameh Kunwar, Dr. Bishnu Prasad Choulagai

Data curation: Kamal Ranabhat

Formal analysis: Kamal Ranabbat, Kiran Shrestha, Ramesh Kunwar

Methodology: Kamal Ranabhat, Dr. Bishnu Prasad Choulagai

Resources: Kamal Ranabhat

Supervision: Dr. Bishnu Prasad Choulagai

Validation: Kamal Ranabhat, Dr. Bishnu Prasad Choulagai

Writing: Kamal Ranabhat, Shubhadra Shahi

